# PROTOCOL FOR THE DEVELOPMENT OF A CORE OUTCOME SET FOR RESPECTFUL MATERNAL AND NEWBORN CARE IN A LOW-MIDDLE INCOME SETTING

**DOI:** 10.1101/2023.05.31.23290715

**Authors:** Farai Marenga, Kushupika Dube, Unice Goshomi, Carol Bedwell, Jamie J Kirkham

## Abstract

**Introduction:** Disrespect and abuse have been seen as a real hindrance to achieving universal coverage for skilled delivery. Improving respectful maternal and newborn care (RMNC) and quality of care around the time of birth has been identified as a key strategy in low- and middle-income countries (LMICs) for reducing the rates of stillbirths and maternal and newborn mortality and morbidity rates (Bohren et al., 2017). Currently, there is no core outcome set (COS) on RMNC, resulting in reporting of various study outcomes from different studies which hinders the improvement of maternal and neonatal health.

**Objective:** To develop a COS for RMNC that can be used in research studies and clinical practice in LMICs.

**Methods /Design:** An exploratory sequential mixed methods evidence synthesis design will be adopted for the study. This design will enable the utilisation of the COS methodology in three stages: (1) A systematic review and secondary analysis of qualitative interviews of women who utilise maternal care services in order to generate a list of outcomes (2) The list of outcomes will be used in a Delphi study with multiple stakeholder groups which include women and their partners, women representative groups, parents, health workers and researchers. Each person will score the outcomes in terms of the defined criteria. (3) The results of the Delphi will be summarised and discussed at a virtual consensus meeting with representation from all stakeholder groups where the final COS will be decided.

**Discussion:** The COS will predominantly be developed for use in a LMIC setting to measure and improve the quality of RMNC services.

## Introduction and Background to the Problem

The World Health Organisation (WHO), United Nations International Children’s Education Fund (UNICEF), (2023) reports that daily approximately 6400 newborns die and about 800 women die due to pregnancy and birth related complications. Furthermore, Sub-Saharan Africa records the highest rates for stillbirths and newborn as well as maternal mortality (545/100000).

Institutional births by skilled health professionals have been identified as a factor that can greatly reduce the maternal and newborn mortality rates (Vogel et al., 2016). However, LMICs’ access to quality health services is not always guaranteed and even when the services are available and accessible the quality may be compromised by disrespect, abuse and mistreatment during childbirth (Shakibazadeh et al., 2018). Disrespect and abuse have been seen as a hindrance to achieving universal coverage for skilled care at birth. Therefore, improving RMNC and quality of care around the time of birth has been identified as a key strategy in most LMICs for reducing stillbirths, newborn and maternal morbidity and mortality. This is because studies have shown that the care a woman receives during pregnancy and childbirth has a huge and lasting impact on the woman’s decision to seek health care in the future (Bohren et al., 2014)

The WHO, (2018), defines Respectful Maternal Care (RMC) as care that is organised for and provided to all women in a manner that ensures that their dignity, privacy and confidentiality is maintained, ensures that they are free from harm and mistreatment, enables informed choice and continuous support during labour and childbirth. The Pan American Health Organisation (PAHO) Virtual Campus (2021), defines Respectful Newborn Care (RNC) as “an approach that focuses on the individual human rights which frame aspects related to ethics, rights and interpersonal relationships, including the respect for women and the newborn fundamental rights, such as autonomy, dignity, decision making and preferences.”

The issue of RMNC has received worldwide attention and several researchers have identified categories and typologies for RMNC as well as disrespect and abuse of women. Bowser and Hill (2010), in their landscape analysis devised categories for disrespect and abuse of women which drew on human rights and ethical principles as, physical abuse, non-consented care, non-confidential care, non-dignified care, discrimination based on specific patient attributes, abandonment of care, and detention in facilities. This typology however, had its limitations thus Bohren et al. (2015), conducted a mixed method systematic review to come up with an evidence based typology which identified seven domains of mistreatment as follows; physical abuse, sexual abuse, verbal abuse, stigma and discrimination, failure to meet professional standards, poor rapport between women and providers and health system conditions and constraints

These typologies have been a basis for formulation of interventions that can be used to reduce and/or eliminate incidences of disrespect and abuse of women thus, promote RMNC. The most common type of intervention for RMC that is implemented especially in LMICs is training of health workers to be advocates for RMNC (Reis et al., 2016).

COS are disease or health care specific and take into account both the potential benefits and harms of interventions (Kirkham et al., 2017). The use of core outcome sets reduces inconsistencies, allowing results from different studies to be compared and combined. It also means research is more likely to report relevant outcomes and reporting bias may be minimised. This is because researchers are expected to report on all the core outcomes or state explicitly why particular outcomes are not reported (Williamson et al., 2012).

The WHO acknowledges that coming up with and choosing the most important outcomes is critical to producing useful guidelines (World Health, 2014). This study aims to develop a COS for RMNC which can be used by researchers in LMIC to report their outcomes.

### Purpose of the study

The purpose of the study is to develop a COS for RMNC that can be used in research studies and clinical practice in LMICs.

### Objectives of the study

1. To systematically review outcomes that are currently being reported in research studies on RMNC.
2. To use qualitative methods to establish the outcomes that are most relevant to key stakeholders of maternal and newborn care.
3. To use consensus-based approaches to have key stakeholders agree on a COS that can be used in future research on RMNC as well as for routine care of women and newborns utilising maternity and newborn care services.
4. To disseminate and implement the COS for RMNC

### Scope of the study

The study aims to develop COS for RMNC. The COS will be developed for research and clinical practice and will consider all interventions and care options for RMNC within this scope. This COS will predominantly be developed for use in an LMIC setting and will be for women who utilise maternal and newborn health services regardless of having had an institutional or home birth.

#### Steering Committee Membership

The Study Steering Committee (SCC) will comprise membership from a multidisciplinary team within a NIHR Global Health Research Unit (GHRU) on the Prevention and Management of Stillbirths and Neonatal Deaths in Sub-Saharan Africa and South Asia. The team includes health care professionals, community engagement and involvement representatives, including women who utilise maternal and newborn health services and methodologists (inclusive of a COS development expert) representing six Sub-Saharan African countries (Kenya, Malawi, Tanzania, Uganda, Zambia and Zimbabwe), two countries in South Asia (India and Pakistan) and experts from the United Kingdom with substantial collaborative research experience in an LMIC setting.

### Existing knowledge of Outcomes

COS development projects relevant to the current study that have been published or ongoing were searched for in May 2022 in the COMET and The CoRe Outcomes in WomeN’s health (CROWN) databases. The studies were searched under disease category “Pregnancy and Childbirth” and disease name, ‘maternal care and neonatal care’. There was no COS for RMC or RMNC that was registered with these databases at the time of the search.

### Methods/Design

The Core Outcome Set-STAndardised Protocol Items: the COS-STAP Statement (Kirkham et al., 2019) was followed for development of this COS protocol. This study is registered on the Core Outcome Measures in Effectiveness Trials (COMET) database https://www.cometinitiative.org/Studies/Details/2100

### Overview of the study design

This study will use the COS development approach guided by the COMET Handbook (Williamson et al., 2017) and the Core Outcome Set-STAndards for Development (COS-STAD) recommendations (Kirkham et al, 2017)and Core Outcome Set–STAndards for Reporting (COS-STAR) Statement as a reporting guideline (Kirkham et al., 2016) will be followed in the development of the COS. An exploratory sequential mixed method design underpinned by the pragmatic philosophical perspective will be adopted for the study. This allows integration of both qualitative and quantitative research, thereby providing a better understanding of what the stakeholders especially the women and parents value and recommend to be included in the COS than either approach alone (Leech and Onwuegbuzie, 2009). This design is also appropriate for the COS development approach which involves three steps shown in Figure 1 below:

**Figure 1:**
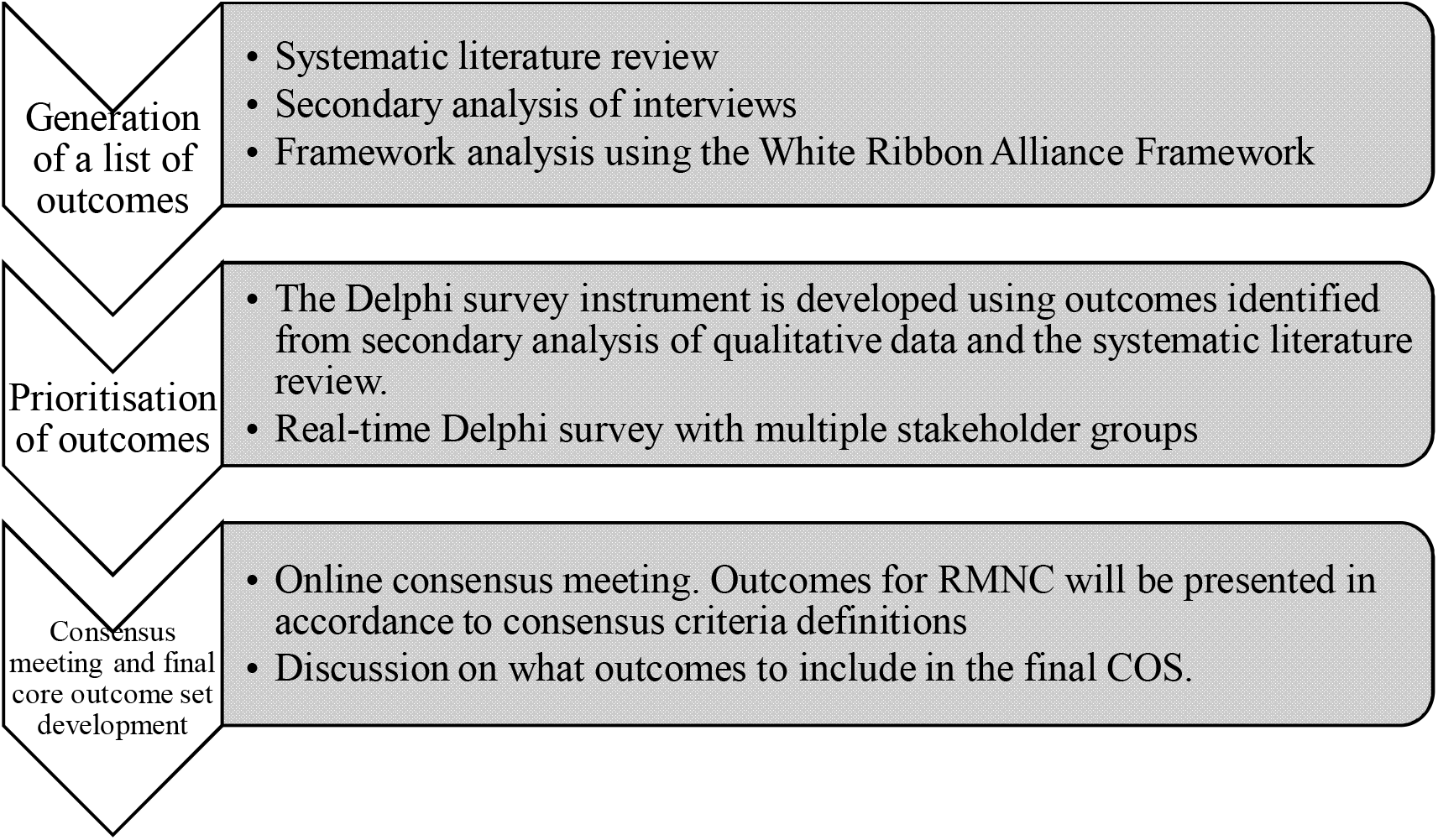
COS Development approach for Developing a COS for RMNC.

## Step 1: Generation of initial lists of outcomes

(1) Generating a list of outcomes by systematically reviewing existing literature from different databases on RMNC and through secondary analysis of qualitative interviews of women who utilise maternal health services, health care professionals as well as researchers working in this field across Sub-Saharan Africa and South Asia.

This step entails a systematic review of literature which will be synthesised with reanalysis of qualitative interviews of the patients Framework analysis will be used to group the outcomes according to their domains.

### a) Systematic Literature review

#### Research Question

Which outcomes and outcome measures are currently being reported in trials and studies for RMNC research?

#### Method

A systematic literature review will be conducted so as to identify and come up with a list of outcomes currently being reported in studies on RMNC. The systematic review protocol for this study was prospectively registered on PROSPERO. Registration number CRD42023354521.

### b) Secondary analysis of interviews

#### Research question

Which outcomes can be identified from the secondary analysis of interviews?

A secondary analysis of interviews done in Malawi, Tanzania and Zambia on RMNC will be undertaken. The NIHR programme of work includes further research projects related to RMNC in these countries. The NIHR unit and the previous NIHR group has considerable data in the form of qualitative interviews conducted in these countries which can be accessed. The aim of this phase of work is to identify any other outcomes from secondary analysis of interviews that were originally completed for other studies on RMNC. Outcomes identified through secondary analysis of interviews and the systematic review will be analysed using the framework analysis method, providing a deductive and inductive approach (Spencer et al., 2003). The framework domains will be derived from the White Ribbon Alliance Charter, with additional domains added as required. These domains include but are not limited to; freedom from harm and ill-treatment, information and informed consent, autonomy and self-determination, privacy and confidentiality, identity and nationality, freedom from discrimination, non-separation of mother and baby adequate nutrition and safe water

## Step 2: Prioritisation of outcomes (Delphi Process)

### Stakeholders

Studies which have involved patients, stakeholders and/or the public have come up with outcomes that may have not been previously identified by other stakeholders (Williamson et al., 2012). Once the outcomes have been identified and listed, stakeholders will be chosen from:

### Community Engagement and Involvement Group (CEI)

The CEI group will include women who are pregnant and those who have given birth within the past year. Their experiences will assist in refining the research question and identifying additional outcomes and prioritising them.

### Health care professionals

Health care professionals involved in the care of women and newborns that is, nurses, nurse/midwives, obstetricians and paediatricians, will be invited to participate in the study.

### Researchers

Researchers involved in different research areas of maternal and child health and core outcome set developers will be invited to participate in the study.

### Recruitment

Participants representing the stakeholders identified above will be invited via email. The invitation will state the rationale for developing a COS for RMNC and how the Real-time Delphi survey works. In order to increase the reach and sample size the participants may be asked to invite other professionals from their contacts who meet the criteria to also participate in the survey.

### Sampling and Sample size

A pragmatic approach will be used to select participants and determine the sample size to represent the health care workers, researchers and academics and women and their partners.

Participants for the Delphi survey will predominantly be chosen from stakeholder groups identified above in Zimbabwe. Input may be sought from health care providers, researchers and academics from other LMICs in the Global Health Research Unit (GHRU), where additional ethical clearance is not required.

### Real-time Delphi survey

The questionnaire will be constructed and managed through Calibrum real-time Delphi software (Calibrum, 2021). The participants will be asked which stakeholder group they belong to (women and/or their partners, health care professionals and researchers). This will enable the collection of stakeholder specific data.

Women and their partners:

- Age,
- Period since using maternity and/or neonatal services,
- Maternal and neonatal health services utilised (public government hospitals, public council clinics, private hospitals/clinics).

Health workers

- Profession (nurse/midwife, obstetrician, paediatrician etc)
- Country of practice
- Organisation affiliation (public government hospitals, public council clinics, private hospitals/clinics/practice).

Researchers

- Country of practice
- Area of research interest
- Organisation affiliation

Stakeholders will be invited to score individual outcomes in a real-time Delphi survey for RMNC. Real-time Delphi surveys have the prospect of improving the speed and efficiency of gathering opinions on outcomes than standard multi-round approaches (Quirke et al., 2021). Participants will be asked to rate the importance of each outcome on a 9-point Likert scale (1-3 limited importance, 4-5 important but not critical and, 7-9 critical) following the GRADE (Grading of Recommendations, Assessment, Development and Evaluations) guidelines (Guyatt et al., 2011). On scoring, participants will immediately receive anonymised feedback which will consist of the individuals rating and the distribution of scores for each outcome according to a) each stakeholder group and b) overall across all stakeholder groups. Once feedback is received, participants will have the opportunity to rereview outcomes and change their scores if they wish. At the end, participants may also add any additional outcomes they think important but not already included in the list. Any additional outcomes will be reviewed by the SCC to consider whether these outcomes are relevant and not duplicated. Participants will be reminded by email to re-visit and re-rate outcomes before the survey ends, where they will also be able to score, receive feedback and re-score the additional outcomes if necessary. The real-time Delphi survey will be managed through Calibrum real-time Delphi software (Calibrum, 2021).

#### Consensus Criteria

On completion of the Delphi survey, the results will be summarised according to the pre-specified definition of consensus (Table 1).

**Table 1:**
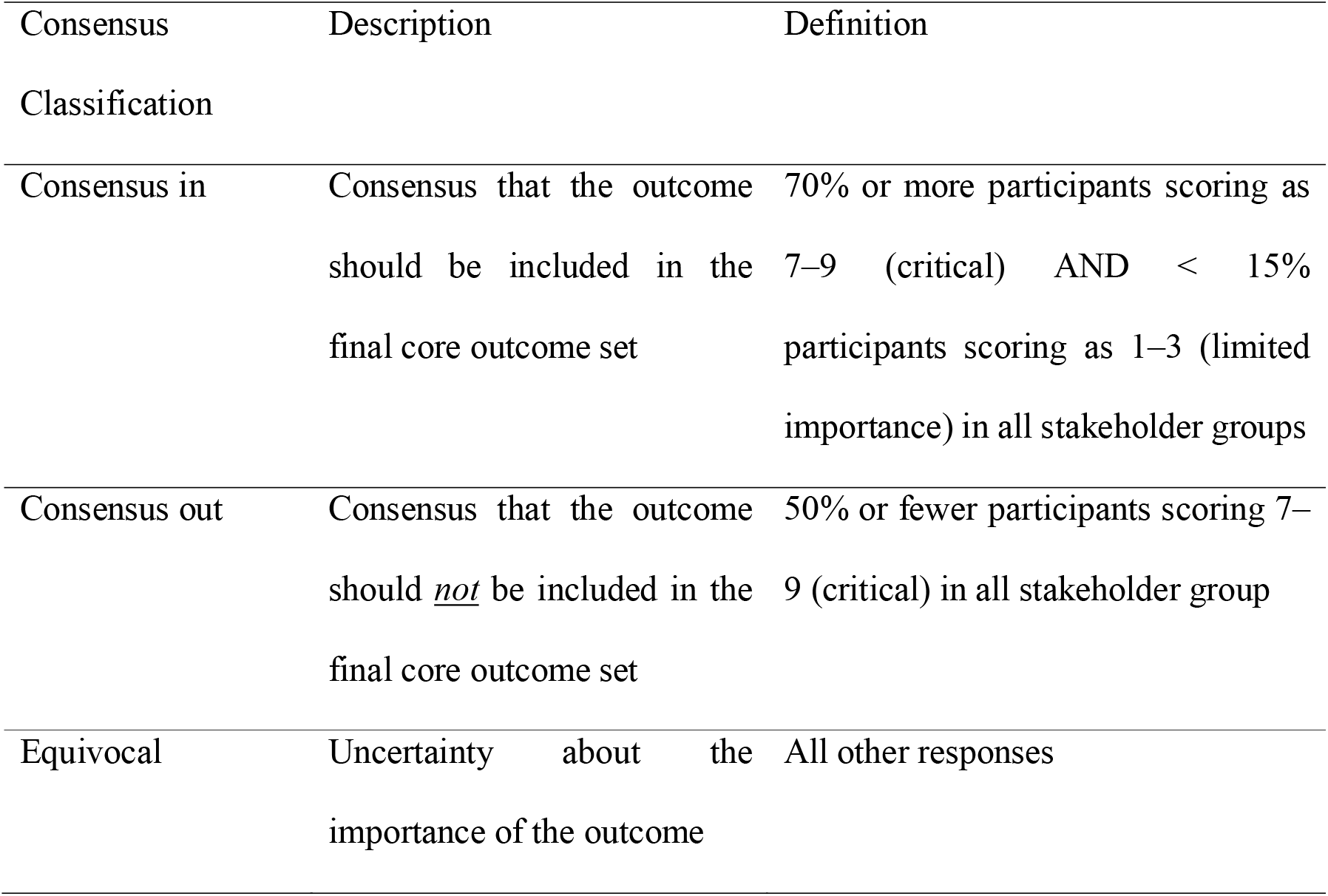
Consensus criteria.

## Step 3: Consensus meeting and final core outcome set development

### Consensus meeting

It is now increasingly well accepted that the future of collaborative and influential research is to bring together diverse key stakeholders especially the patients to reach a consensus (Williamson et al., 2012). The consensus meeting will be done online. The results to all outcomes for RMNC will be presented in accordance to consensus criteria definitions (Table 1) from the Delphi survey. Where outcomes from the Delphi reached ‘consensus in’ or ‘consensus out’, participants will be invited to briefly discuss and provide more information if they disagree with the inclusion/exclusion of the outcome in the COS. Following discussion, participants of the consensus meeting will re-score the outcome. Where outcomes were equivocal during the Delphi survey, they will be discussed and participants of the consensus meeting will be invited to re-score the outcome. Where voting is required, this will be undertaken anonymously using the same criteria and consensus definition as used in the Delphi survey (Table 1).

### Final core outcome set

At the end of the consensus meeting, the consensus meeting panel will review the proposed outcomes to be included in the COS following the discussions and voting. A final reflective discussion will be undertaken to ensure the outcomes included are pragmatic and feasible to measure in an LMIC setting. If a final COS is not agreed on at the end of the consensus meeting, subsequent online meetings will be considered in order to ratify the final COS.

## Ethical and legal considerations

Ethical approval will be sought from the Research Ethics Committee at Women’s University in Africa before the study is commenced. Participant consent will be obtained in verbal (where possible) and then in written form. Ethical principles including voluntary participation, ensuring participant’s right to privacy, anonymity and self-determination will be observed during the study and thereafter. Study participants will to the best of the researcher’s ability be protected from any harm physically and psychologically.

## Discussion

Development of COS is essential in improving health care and reduce research waste. This will be the first COS to be developed for RMNC and will be predominantly used in the LMIC setting. It will be of particular importance in the LMICs where disrespect and abuse are more prevalent and where the highest burden of maternal mortalities (Kassa et al., 2020)

The inclusion of outcomes derived from secondary analysis of interviews will enable the capturing of outcomes from the patients’ perspective. Until recently very few COS studies originated from LMIC and the development of COS did not include participants from the LMIC setting. This led to development of COS which include outcomes which may not be measurable in these settings (Karumbi et al., 2021).

This study will be part of the few studies on development of COS originating from LMICs, therefore it will help to understand the challenges that researchers who develop COS in LMICs face and how they can be mitigated. This information will help formulate and implement robust methodology in developing COS, ways to disseminate and implement in LMICs thus, ensuring the uptake and use of COS in these settings.

## Data Availability

All data produced in the present study are available upon reasonable request to the authors

https://www.cometinitiative.org/Studies/Details/2100

## Acknowledgements

We would like to acknowledge the contributions from colleagues from the World Health Organisation, Portela A.G and Tuncalp O.

